# The Effectiveness of Social Distancing in Mitigating COVID-19 Spread: a modelling analysis

**DOI:** 10.1101/2020.03.20.20040055

**Authors:** George J Milne, Simon Xie

## Abstract

**Background:** The novel coronavirus COVID-19 has been classified by the World Health Organisation as a pandemic due to its worldwide spread. The ability of countries to contain and control transmission is critical in the absence of a vaccine. We evaluated a range of social distancing measures to determine which strategies are most effective in reducing the peak daily infection rate, and consequential pressure on the health care system.

**Methods:** Using COVID-19 transmission data from the outbreak source in Hubei Province, China, collected prior to activation of containment measures, we adapted an established individual based simulation model of the city of Newcastle, Australia, population 272,409. Simulation of virus transmission in this community model without interventions provided a baseline from which to compare alternative social distancing strategies. The infection history of each individual was determined, as was the time infected. From this model-generated data, the rate of growth in cases, the magnitude of the epidemic peak, and the outbreak duration were obtained.

**Findings:** The application of all four social distancing interventions: school closure, workplace non-attendance, increased case isolation, and community contact reduction is highly effective in flattening the epidemic curve, reducing the maximum daily case numbers, and lengthening outbreak duration. These were also found to be effective even after 10 weeks delay from index case arrivals. The most effective single intervention was found to be increasing case isolation, to 100% of children and 90% of adults.

**Interpretation:** As strong social distancing intervention strategies had the most effect in reducing the epidemic peak, this strategy may be considered when weaker strategies are first tried and found to be less effective. Questions arise as to the duration of strong social distancing measures, given they are highly disruptive to society. Tradeoffs may need to be made between the effectiveness of social distancing strategies and population willingness to adhere to them.

## Introduction

At the early stages of the COVID-19 coronavirus pandemic originating in Wuhan, China data on virus transmissibility and pathogenesis was uncertain, as would be the case for a novel influenza virus.^1^ Given this uncertainty, Chinese authorities adopted strict measures to contain COVID-19 spread, by continuing closure of schools and workplaces, which were already closed for the Chinese Lunar New Year, activating measures to enforce significant community contact reduction, and closing transport links between population centres. This response was in contrast to the situation which occurred in 2002/3 with the Severe Acute Respiratory Syndrome epidemic, as with the current coronavirus also originating from an animal reservoir in China, where the SARS coronavirus had spread widely before measures were effected to contain it.^2^ Given the resulting worldwide spread of COVID-19 public health authorities require guidance on how best to mitigate its spread, to reduce the peak in daily case numbers, and so lessen the number of critically ill cases requiring hospitalization. We present results from an extensive model-based analysis of the effectiveness of social distancing interventions, the only measures currently available in the absence of vaccines and antiviral drug treatments, to help inform health authorities.

Coronaviruses are respiratory viruses where transmission between individuals occurs primarily via aerosol droplets. Close contact between individuals at home, in schools and workplaces, on transport, and at community gatherings is necessary for virus transmission to occur. Methods to reduce this contact, namely social distancing, were enacted in urban centres with diagnosed COVID-19 cases, as in Hubei Province, China in January 2020.

As of early March 2020 many key characteristics of the novel coronavirus are still uncertain, though some consensus is beginning to emerge.^3^ Key characteristics are its transmissibility, denoted by its *basic reproduction number R***0**, the average number of secondary cases caused by one infected individual in an otherwise uninfected population, and its severity, represented by the *case/fatality ratio* (CFR).^4^ The reproduction number is a key metric; it helps determine whether a pathogen introduced into a community will spread and, significantly, gives guidance as to its rate of spread. The aim of containment measures is to reduce the reproduction number to below 1.0 when the outbreak will eventually fade out.

Social distancing measures aim to prevent onward person-to-person virus transmission by minimizing contact, measures that are currently in place in China, Northern Italy, South Korea and increasingly in other parts of the world.

Using initial transmission data from the COVID-19 epidemic, we adjusted parameters in an established, individual-based simulation model of an Australian community to reflect these COVID-19 characteristics, and applied the model to evaluate the effectiveness of a range of social distancing interventions. We also considered these mitigation measures under a higher transmission “worst-case” scenario.

The results, in terms of a reduction in the number of cases and the rate of growth in case numbers, provide guidance to public health authorities as to how to optimise containment and control measures. Key questions that health authorities require guidance on involve the magnitude of social distancing interventions required to arrest virus spread, and include the strength, compliance rate, and duration of control measures required to be effective.

Containment and control is reliant on social distancing measures and the focus of this study is on measures to mitigate virus transmission within communities, whether towns or large cities. Halting movement between cities and between countries, as has already occurred with the COVID-19 pandemic, is an additional measure to adopt in any pandemic situation; others have examined these control measures for COVID-19 and pandemic influenza settings.^5,6^

## Methods

A community-based simulation model capturing the demographics and movement patterns of individuals in an Australian city, with the virus transmission characteristics of COVID-19, was used to evaluate the potential effectiveness of a range of social distancing strategies. The model is individual-based (*c*.*f*. agent-based) and represent each individual in a specific community, matching recent census and other government data.^7,8^ We previously developed individual-based simulation models for population centres in Australia, South Africa, Thailand, Vietnam and Papua New Guinea, all using the same underlying automata-theoretic modelling methodology, to capture the dynamics of both pathogen transmission and population mobility.^9-13^ This modelling methodology is presented in detail in previous publications.^11^

This approach to disease modelling allows us to explicitly simulate person-to-person virus transmission, the probability of such transmission, the location of transmission (*e*.*g*. school, workplace, home, community) and determines each individual’s infection status through time.^10-12,14^ Simulation models create a “virtual world” of individuals whose daily movement, changing contact patterns and disease biology dynamics aim to replicate that of the real-world system in as much detail as data sources permit, such as data from the POLYMOD contact pattern study.^15^

We modelled an Australian city, Newcastle in New South Wales. Its model matches the real-world counterpart with respect to population size, household structure, age of individuals in each household (stratified from Australian census data into ten age bands), employment, schooling, and daily movement between these locations. The model was developed using detailed census, workplace and mobility data using a model development methodology applied previously.^11^ Such models create realistic representations of the respective communities at an individual-by-individual level using the best available data sources, including from the Australian Bureau of Statistics (ABS).^16^

The Newcastle model represents 272,409 people in an urban area with a population which is representative of the Australian population as a whole, in terms of age distribution. ABS census data^7,8^ were used to capture the age-specific demographics of every household in the community. Data for schools, including geographical location and pre-primary, primary and secondary enrolment numbers for each school in the Newcastle were obtained from New South Wales state government publications.^17^ ABS data were also used to determine workplace locations and workforce sizes. These data were used to generate a model which captures the movement and contact patterns of individuals on a day-by-day basis.^15^

Simulation model parameter settings to reflect the transmission characteristics of the COVID-19 epidemic were determined via calibration to represent an unmitigated outbreak with a basic reproduction number R_0_ of approximately 2·2, taken from work by Li and colleagues.^3^ This basic reproduction number corresponds to that derived by Kucharski and colleagues.^18^ These data provided virus transmission settings for the model corresponding to the spread characteristics in Wuhan, China prior to activation of social distancing measures (which started on 23^rd^ January 2020). These parameter settings gave us an unmitigated epidemic baseline from which to compare alternative social distancing (SD) strategies. Model outputs obtained by running the simulation software for the duration of an infectious disease outbreak gave the infection history of each individual in the modelled community. This data was used to determine the total number of infectious individuals, where and when infection occurs, and may be used to determine the resulting health burden in terms of hospitalisations and deaths.^14,19^ The data generated by running the simulation model software provided data on the rate of growth in cases, the magnitude of the peak number of cases and resulting impact on the hospital system, the time to reach the peak, and duration of the outbreak.

To determine the effectiveness of specific social distancing interventions we compared the number of infectious cases generated by simulating the unmitigated COVID-19 outbreak with one which has specific social distancing measures in place. A series of modelling experiments were conducted with alternative social distancing strategies activated, giving the infection dynamics of the modelled community with a specific mitigation strategy in place. The difference in cases allows for quantification of intervention effectiveness, and the benefit represented as a reduction in infections and thus symptomatic cases and deaths.

A transmission parameter is used to model the probability of COVID-19 transmission following contact between a susceptible and an infectious individual. That is, a pairing of an individual in infectious state *I* and one in susceptible state *S*, as in the *S-E-I-R* state transition representation of the spread dynamics of a virus.^4^ Adjusting this transmission parameter allowed us to replicated epidemics with different reproduction numbers, and thus attack rates.

We have assumed the transmission characteristics of COVID-19 using early data from the outbreak in Wuhan, prior to social distancing interventions.^3^ These are as follows: a basic reproduction number R_0_ of 2·2; an incubation period averaging 5·5 days, from infection to symptom emergence (if any); a latent period averaging 4·5 days, from infection to becoming infectious; an infectious period averaging 3.0 days, the first day being asymptomatic; and a 20% asymptomatic rate, that is, 20% of infectious individuals show no symptoms and are not classed as cases. These data are contained in Table 1. At the early stage of the COVID-19 epidemic reliable age-specific symptomatic attack rates were unavailable and we thus assumed transmission between different ages of individuals occurs similarly. In addition, we evaluated social distancing effectiveness with a higher asymptomatic setting, of 35%.

**Table 1:** Summary of simulation parameters

| Parameter | Value |
| --- | --- |
| **Baseline Asymptomatic Ratio** | 20% |
| **Targeted $R_0$** | 2.2 |
| **Simulation Output Infection Rate** | 66.00% |
| **Infection** |  |
| **Latent Period (Exposed)** | 4.5 days |
| **Incubation Period (Including Latent)** | 5.5 days |
| **Infectious period** | 2 days (Finish at day 7.5) |
| **Post-symptomatic period** | None |

The model explicitly represents each household, workplace and school in community and the movement of individuals between, as they move from households to schools and workplaces in a daytime cycle, then return to their household in an evening cycle, with each day split into a day and night period. This mobility mechanism allowed us to model changing contact patterns, with possible virus transmission occurring in these contact locations, and also in the wider community, including at weekends. The model represents the individual-to-individual contact patterns in as much detail as data sources provide, to accurately describe how the movement of individuals allows virus transmission to spread over a geographic region. This level of detail is critical for modelling social distancing interventions, whose aim is to minimize person-to-person contact patterns and consequential virus transmission. Such models have been used previously to evaluate the effectiveness (and cost-effectiveness) of alternative infectious disease control and containment strategies, including social distancing, vaccination and antiviral drug treatment in an Australian setting.^19^

### Social distancing

Four distinct social distancing measures are available to health authorities: *a*) school closure; *b*) workplace closure and non-attendance; *b*) case isolation; *d*) reduced community-wide contact.

Assumptions regarding feasible social distancing measures made in prior pandemic influenza modelling studies are also applicable in the novel coronavirus context.^2,3^ These may be briefly explained as follows. We consider both moderate workplace and community contact reductions, as well as much higher reductions for these two mitigation strategies. School closure: when schools and further education institutions are closed students have contact with household members during the daytime and also have contact in the community. Workplace non-attendance: 50% of all persons in the workforce are absent, have contact in the home in the daytime and still having contact in the wider community. Increased home isolation of cases: 90% of adults and 100% of children withdraw to the home on becoming ill *i*.*e*. are symptomatic and only have contact with household members; this is an increase from the baseline assumption of 50% adults and 90% children withdrawal due to illness. Community contact reduction: contact in the wider community is reduced by 30%.

We also evaluated strengthening these measures, by increasing workplace non-attendance to 90% and reducing community contact by 50% or 70%. These higher contact reductions may correspond to the social distancing measures taken in Hubei Province, China. To provide information on when social distancing measures should be activated, we further analysed the impact which delayed activation has on the epidemic growth rate.

While the aim of vaccination is to reduce the reproduction number *R*_0_ of an outbreak to less than 1.0 and an outbreak will eventually fade out, the situation with COVID-19 is different. As with the SARS outbreak in 2003 no vaccine is available,^20^ and the population has no immunity at the outset. Reliance must be made on robust social distancing interventions, contact tracing and early isolation of diagnosed cases. The aim of social distancing interventions in the current situation is to slow down transmission and reduce the growth rate in case numbers. This approach aims to lessen the daily pressure on health care personnel and hospital facilities, such as intensive care beds, and to lower mortality rates.

Using the infection characteristics of COVID-19 in Table 1, we conducted a range of simulation experiments. These involved 9 alternative social distancing mitigation strategies interventions and 11 activation delays, between zero to ten weeks.

## Results

The highest reduction in the infection attack rate is achieved by the rapid activation of all available social distancing interventions, and with the highest rates of compliance. With an activation delay of up to six weeks from arrival of the first infectious cases into the modelled community, the continued use of all four social distancing interventions with 90% workplace non-attendance and a 70% reduction in community-wide contact resulted in a reduction of the infection rate from 66% to less than 1%, see Table 2. Similarly, all four social distancing interventions with lower workplace non-attendance (50%) and a lower reduction in community-wide contact (by 30%) and activation delays of up to ten weeks also held the infection rate to below 10%. With these “very high” and “high” social distancing measures activated an outbreak can be substantially contained.

**Table 2:** Total infection rate for alternative social distancing intervention and increasing activation delays.

| Interventions | N/A (Baseline) | School Closure (SC) | Increase d Case Isolation (CI) | Workplac e Non-attendance 50% (WN50) | Communit y Contact Reduction 30% (CCR30) | CCR50 | All together (SC+ CI+ WN50+ CCR30) | WN90 | CCR70 | All (SC+ CI+ WN90+ CCR70) |
| --- | --- | --- | --- | --- | --- | --- | --- | --- | --- | --- |
| 0 weeks | 66.00% | 61.90% | 31.02% | 58.77% | 53.45% | 41.54% | 1.14% | 48.89% | 23.67 % | 0.42% |
| 1 weeks | 66.00% | 61.92% | 31.26% | 58.75% | 53.47% | 41.57% | 1.11% | 48.93% | 23.47 % | 0.42% |
| 2 weeks | 66.00% | 61.82% | 31.21% | 58.80% | 53.36% | 41.66% | 1.15% | 48.86% | 23.77 % | 0.43% |
| 3 weeks | 66.00% | 61.88% | 31.25% | 58.70% | 53.42% | 41.40% | 1.21% | 48.86% | 24.13 % | 0.46% |
| 4 weeks | 66.00% | 61.93% | 31.47% | 58.71% | 53.42% | 41.59% | 1.34% | 48.75% | 24.10 % | 0.52% |
| 5 weeks | 66.00% | 61.77% | 31.69% | 58.69% | 53.44% | 41.44% | 1.61% | 48.97% | 24.60 % | 0.64% |
| 6 weeks | 66.00% | 61.87% | 32.01% | 58.69% | 53.47% | 41.63% | 2.11% | 48.91% | 24.83 % | 0.84% |
| 7 weeks | 66.00% | 61.93% | 32.32% | 58.74% | 53.65% | 41.78% | 2.87% | 48.93% | 25.65 % | 1.21% |
| 8 weeks | 66.00% | 61.92% | 32.94% | 58.88% | 53.53% | 42.05% | 4.33% | 49.11% | 26.31 % | 1.85% |
| 9 weeks | 66.00% | 61.97% | 33.90% | 58.84% | 53.73% | 42.38% | 6.49% | 49.36% | 27.07 % | 2.95% |
| 10 weeks | 66.00% | 62.06% | 35.47% | 59.00% | 53.93% | 43.01% | 9.51% | 49.64% | 28.72 % | 4.76% |

For the city of Newcastle, Australia, with a population of 272,409 it is shown that no single social distancing intervention has a significant effect on reducing the overall number of infections, see Table 3. With no significant activation delay, the most effective single measure is the 70% reduction in community-wide contact. This reduces the infection attack rate to approximately a third, from 180,000 to 64,000 (Table 3) and from 66% of the population down to 24% (Table 2). Lower rates of community contact reduction, by 50% and 30% are found to be significantly less effective. The second most effective single measure in increased case isolation, with 100% children and 90% adult compliance. This is found to reduce the overall number of infections from 180,000 to 85,000 (Table 3) and from 66% of the population to 31% (Table 2).

**Table 3:** Infected case numbers for alternative social distancing interventions and increased activation delays, for Newcastle population of 272,409.

| Intervention<br>s | N/A<br>(Baseline<br>) | School<br>Closure<br>(SC) | Increase<br>d Case<br>Isolation<br>(CI) | Workplac<br>e Non-<br>attendance<br>50%<br>(WN50) | Communit<br>y Contact<br>Reduction<br>30%<br>(CCR30) | CCR50 | All<br>together<br>(SC+<br>CI+<br>WN50+<br>CCR30) | WN90 | CCR7<br>0 | All<br>(SC+<br>CI+<br>WN90+<br>CCR70<br>) |
| --- | --- | --- | --- | --- | --- | --- | --- | --- | --- | --- |
| 0 weeks | 179,788 | 168,611 | 84,508 | 160,085 | 145,592 | 113,149 | 3,094 | 133,181 | 64,492 | 1,131 |
| 1 weeks | 179,788 | 168,668 | 85,167 | 160,044 | 145,647 | 113,230 | 3,015 | 133,283 | 63,924 | 1,151 |
| 2 weeks | 179,788 | 168,395 | 85,006 | 160,182 | 145,364 | 113,486 | 3,140 | 133,109 | 64,750 | 1,166 |
| 3 weeks | 179,788 | 168,573 | 85,126 | 159,901 | 145,524 | 112,777 | 3,302 | 133,100 | 65,739 | 1,255 |
| 4 weeks | 179,788 | 168,705 | 85,720 | 159,940 | 145,529 | 113,303 | 3,642 | 132,794 | 65,641 | 1,414 |
| 5 weeks | 179,788 | 168,258 | 86,330 | 159,874 | 145,586 | 112,895 | 4,384 | 133,407 | 67,015 | 1,730 |
| 6 weeks | 179,788 | 168,552 | 87,207 | 159,877 | 145,647 | 113,405 | 5,735 | 133,247 | 67,646 | 2,299 |
| 7 weeks | 179,788 | 168,703 | 88,052 | 160,008 | 146,137 | 113,817 | 7,811 | 133,278 | 69,866 | 3,309 |
| 8 weeks | 179,788 | 168,667 | 89,727 | 160,389 | 145,808 | 114,553 | 11,800 | 133,777 | 71,664 | 5,034 |
| 9 weeks | 179,788 | 168,820 | 92,342 | 160,285 | 146,372 | 115,440 | 17,672 | 134,473 | 73,736 | 8,041 |
| 10 weeks | 179,788 | 169,063 | 96,614 | 160,728 | 146,912 | 117,170 | 25,904 | 135,223 | 78,223 | 12,966 |

Combined social distancing measures were found to be highly effective. With all measures activated simultaneously, and 50% workplace non-attendance and a 30% reduction in community contact, a local epidemic can be halted. Even with an activation delay of eight weeks this combined strategy reduces the overall number of infections from 180,000 to 12,000. With a more rigorous intervention strategy involving school closure, increased case isolation, 90% workplace non-attendance and a 70% reduction in community-wide contact an even greater reduction may be achieved. With an activation delay of up to seven weeks, an outbreak can be effectively stopped, with only 3,300 resulting infections.

The ability to significantly reduce transmission by combining rigorous social distancing interventions together, even applying them after a significant period beyond the initial arrival of infectious cases into the community, is illustrated in Figure 1. Here, the simulation results have been scaled to Perth, Western Australia, which has a population of approximately 2·2 million. This figure presents results from the simulation model assuming a ten week delay in activating social distancing measures following arrival of initial cases into the community. It may be seen that the two combined measures (in green) are able to significantly reduce the daily number of cases, with the more robust intervention (dashed green) being the most effective. Nevertheless, all single measures are seen to reduce the epidemic peak, with all school and further education establishment closure being the least effective and the 70% reduction in community-wide contact the most effective single social distancing measure.

**Figure 1:**
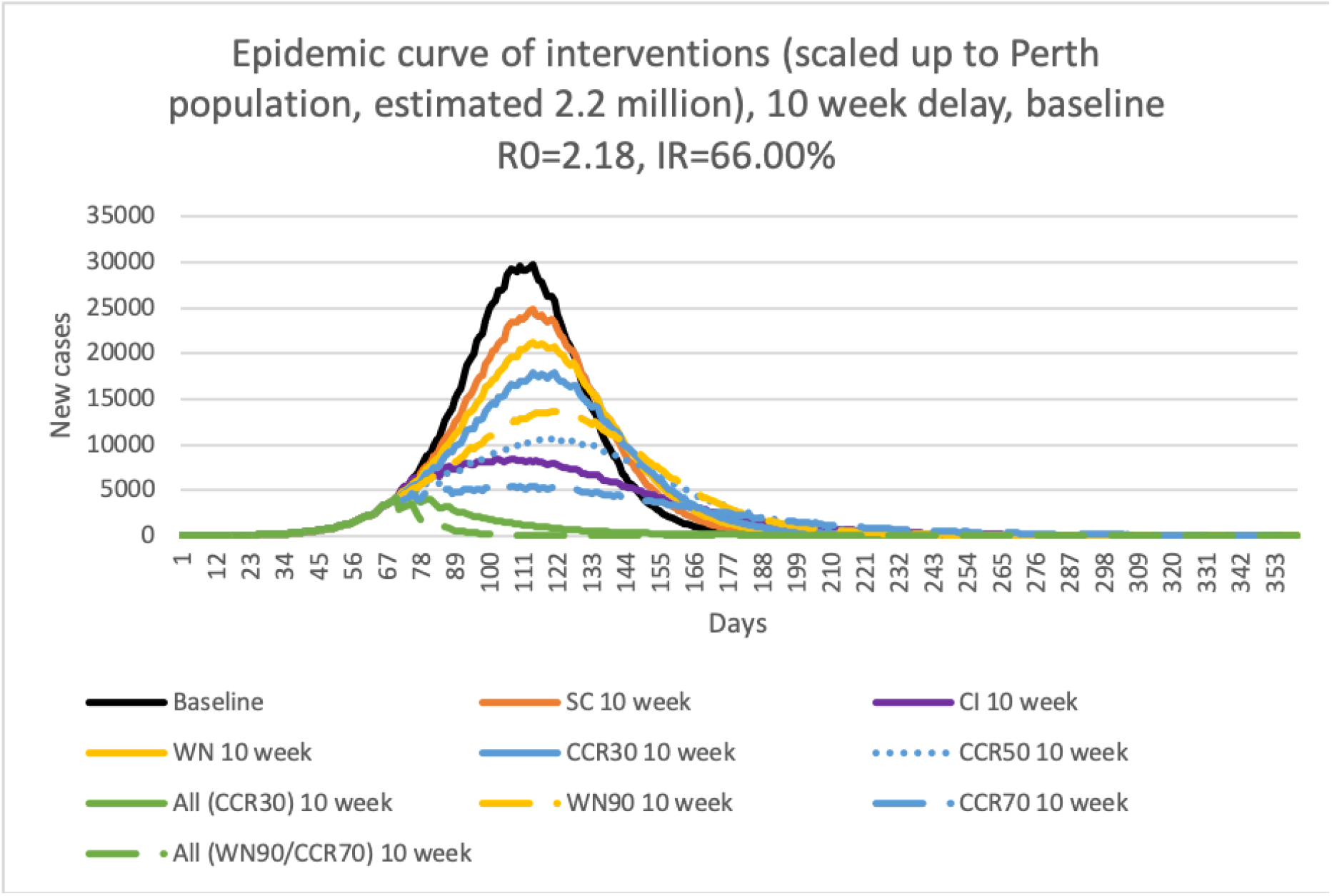
Ten week delay epidemic curve for Perth.

Figure 1 illustrates the growth in cases prior to intervention activation (the increasing green curve) and how the activation of the robust combined strategies at week ten rapidly reduces the number of daily cases, from approximately day 79 onwards. All individual interventions are seen to lower the peak daily number of cases, with a 70% reduction in community cases (CCR70, long dashed blue) being the most effective in reducing the peak, followed by increased case isolation (CI, purple). A less “strict” community contact reduction of 50% (CCR50, dotted blue), 90% workplace absenteeism (WN90, dashed yellow), and 30% community contact reduction (CCT30, solid blue) are single interventions which are increasingly less effective in reducing the daily infection peak.

Figures 2 and 3 present the epidemic curves for infectious cases with one and five week activation delays, respectively. For these short activation delay scenarios both combined intervention strategies are effective in preventing an epidemic occurring, both green lines are flat. However, this assumes that these robust intervention strategies are continued indefinitely. 70% community-wide contact reduction is the most effective single intervention measure and increasing case isolation and a 50% reduction in community contact are also seen to be highly effective in reducing the growth in case numbers and delaying the epidemic peak.

**Figure 2:**
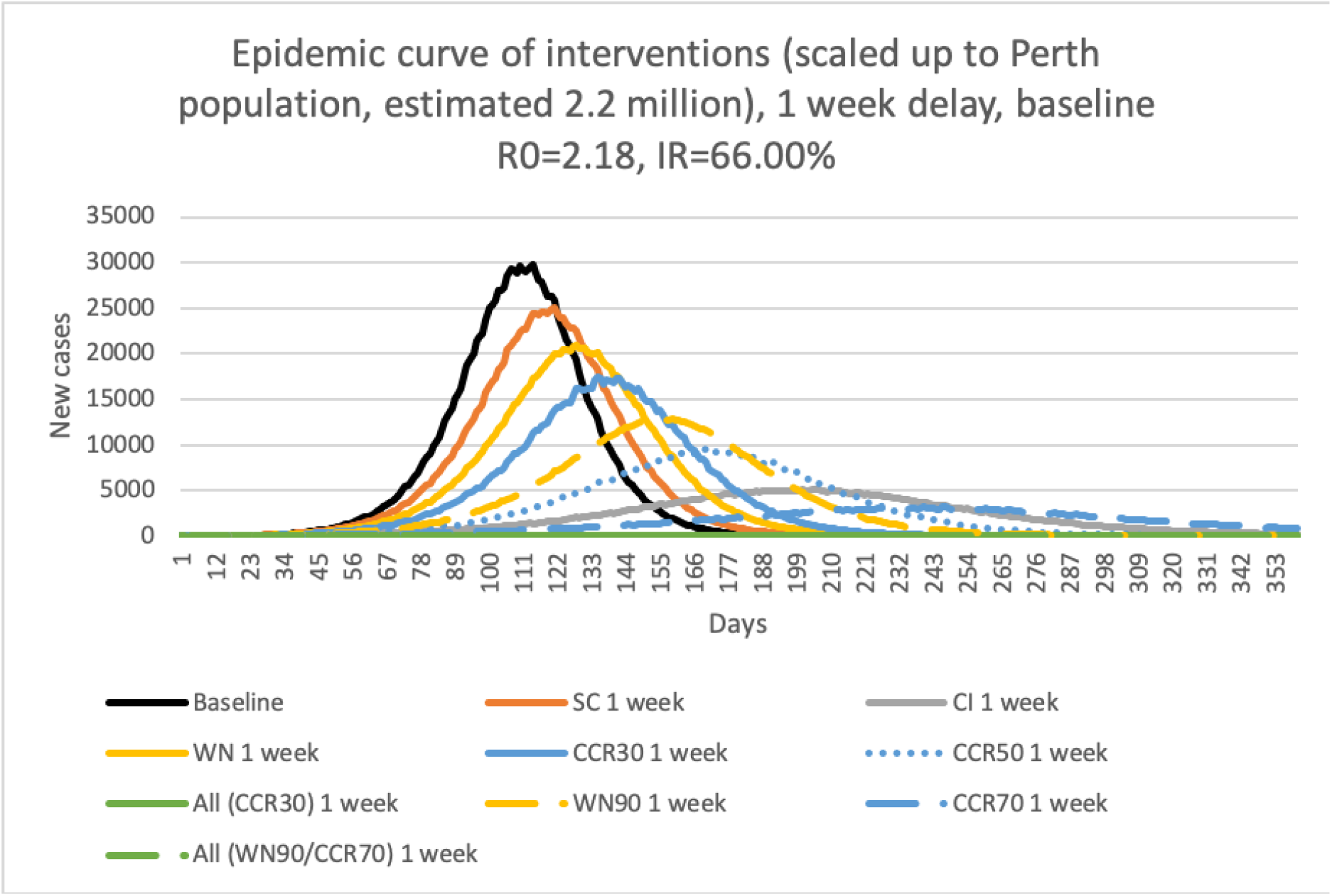
One week delay epidemic curve for Perth.

**Figure 3:**
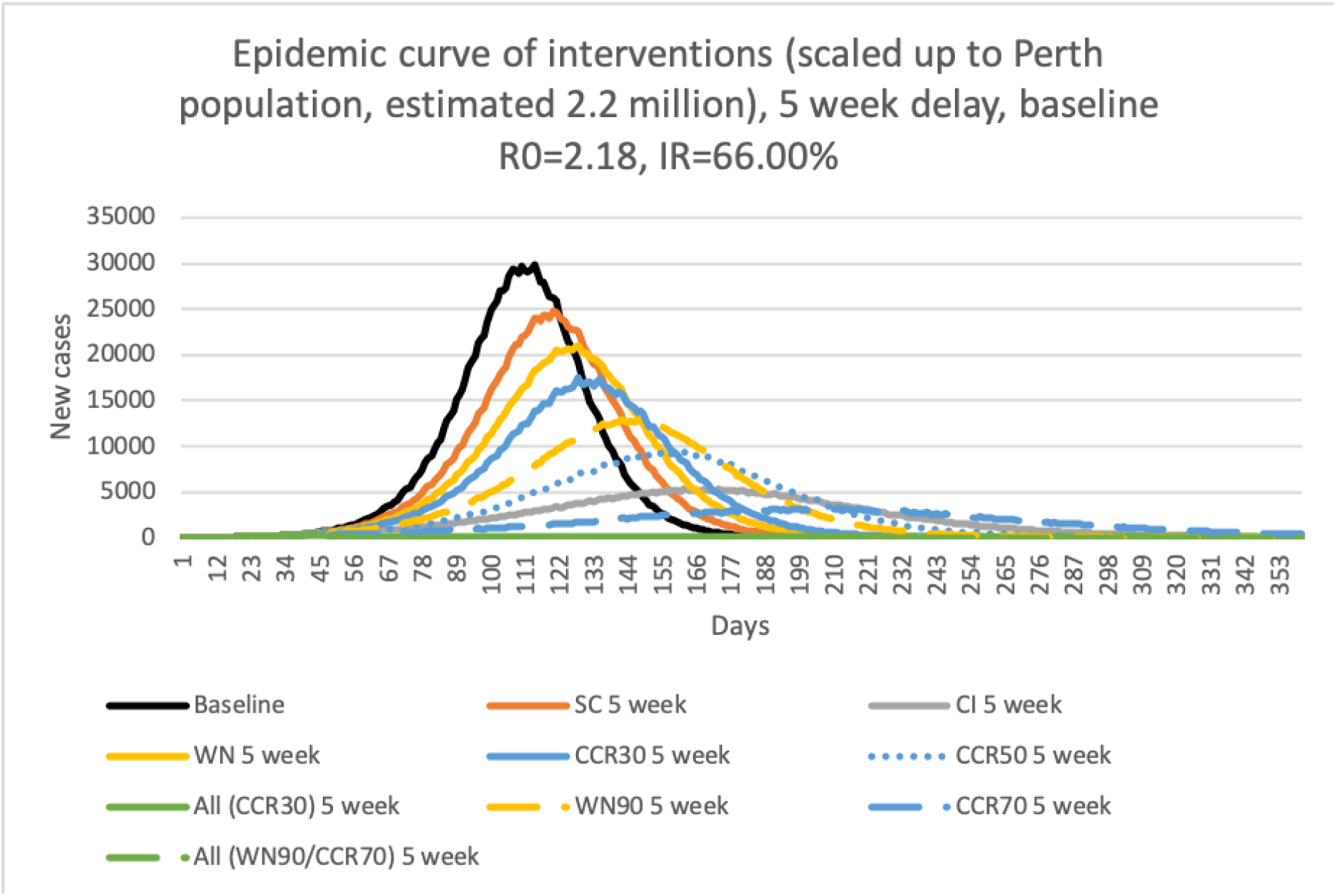
Five week delay epidemic curve for Perth.

Given the vulnerabilty of those aged 65 and above to poor outcomes following COVID-19 infections,^21^ we were able to predict the effectiveness of social distancing interventions on reducing infections in this age group. Figure 4 extracts the daily elderly case infection rate from the same model-generated infection dataset used in Figure 2, that is with a 1 week delay on social distancing activation.

**Figure 4:**
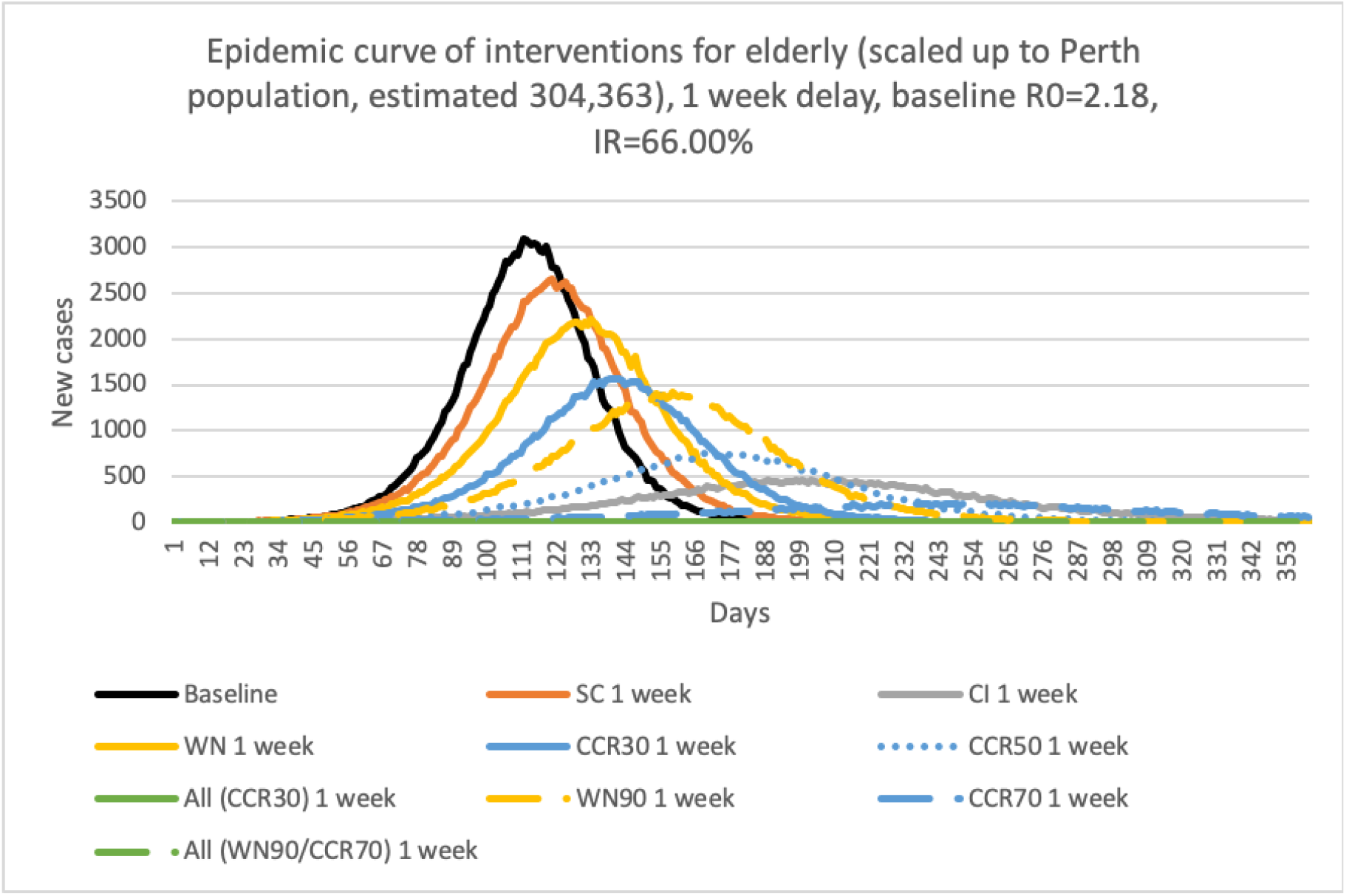
Epidemic curve of infected elderly, scaled to Perth elderly population of ∼304,363.

Figure 4 demonstrates how robust social distancing combining all social distancing measures (green lines) may reduce infections in the elderly to almost zero. A 70% reduction in community contact (dashed blue line) is a single intervention measure predicted to reduce the maximum daily infections in this age group to ∼200, from ∼3000 in an unmitigated scenario. A 50% community contact reduction (blue dots) and increased case isolation (grey) are also seen to be effective in reducing the peak in cases and flattening and lengthening the epidemic curve.

These data are available to predict demand on a country’s health care system. Using COVID-19 data from China,^21^ these elderly infection rates may be used to estimate the daily demand for critical care. As an example, the daily peak in the 70% community contact reduction intervention of ∼200 elderly individuals includes both symptomatic and asymptomatic cases, this may reduce to the order of 150 ill elderly. Assuming approximately 10% of these require critical care,^21^ then a maximum of 15 new cases a day are likely to require treatment in intensive care units (ICUs). Using similar assumptions, the unmitigated peak in elderly cases (black line) is predicted to result in a peak daily demand for ICU places of approximately 225.

The above results on the effectiveness of social distancing measures have assumed that these measures can be held for as long as a year. Figure 5 illustrates a more complex COVID-19 control scenario involving starting and stopping social distancing. Here we assumed a 10 week delay in activation, as in the Figure 1 analysis, and considered two combined strategies which were halted after 6 or 8 weeks repectively.

**Figure 5:**
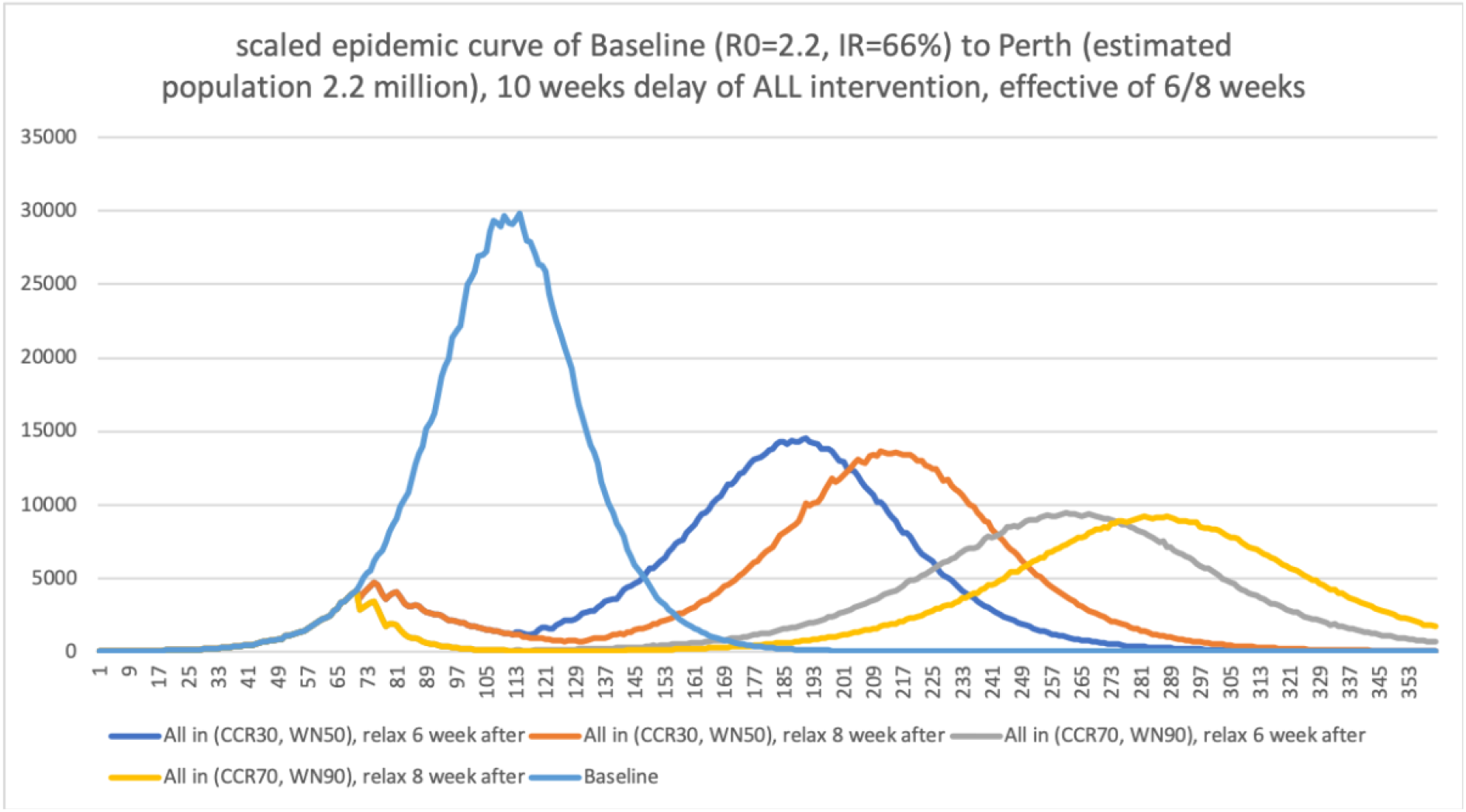
COVID-19 predicted epidemic curve for overall population of Perth.

The two strategies illustrated in Figure 5 both involve increasing case isolation to 100% of children and 90% of adults, and closing all schools. In addition, one involves at 30% reduction in community contact and a 50% reduction in workplace attendance (CCR30, WN50) as in the above figure. The other is a more robust scenario, with community contact reduction strengthened to a 70% reduction, and workplace closure resulting in a 90% non-attendance rate (CCR70, WN90). We consider halting these robust, combined interventions after 6 weeks or 8 weeks.

The four scenarios, all with a 10 week delay in activation, can be seen to significantly lessen daily case numbers in the initial phase, reducing them rapidly from around day 73. Note that the orange and yellow curves overlay the dark blue and grey curves at first, from week 11. The pale blue curve indicates the pattern of daily cases with no social distancing in place, involving both symptomatic and asymptomatic cases.

All four scenarios predict a rapid gain in daily cases after interventions cease, 6 or 8 weeks after activation, under strong (CCR reduced by 30% and WN by 50%) and very strong (CCR reduced by 70% and WN by 90) social distancing. It can be seen that case numbers increase again, as we would expect, with the very strong interventions (grey and yellow) having a flatter and longer epidemic curve.

## Discussion

It is apparent from results generated by our simulation model that both the timing and strength of social distancing measures have a substantial effect in reducing the number of infected individuals in a pandemic situation. In reality, it is unlikely that the initial arrival of infectious cases into a community would be identified in a matter of a few days. This suggests that delays may be expected if waiting for diagnosis to occur before activation of mitigating measures. The modelling results suggest that even with a significant delay in invoking mitigation strategies, 10 weeks in the case of data presented in Figure 1, this delay and consequential growth in case numbers may be countered by the scale of interventions adopted, by combining multiple robust social distancing measures.

The timing of activation of social distancing measures is a challenge facing public health authorities, balancing what needs to be done with what is feasible, and this will vary between countries. Our modelling gives initial guidance on the relative benefit of a range of mitigation strategies. As the COVID-19 pandemic develops more subtle strategies will need to be evaluated, such as the phased introduction of additional measures if it is found that existing strategies are ineffective in reducing daily case numbers. Similarly, modelling will be required to determine optimal strategies to phase the ending of interventions once the epidemic peak has passed. Models such as that presented here will have a key role in analysing these evolving situations.

The results indicate that two separate social distancing measures are highly effective, case isolation and a 70% reduction in community-wide contact. Both of these measures may be strengthened further. Given we assumed that only cases are isolated, not the whole family, there is scope to increase the effectiveness of that strategy. The 70% community contact reduction intervention may also be further strengthened to a 90% reduction if required.

Deciding on the strength or robustness of interventions will be a challenge for governments. There will be a need to balance what may be necessary to reduce the daily infection rate, and take pressure off health care resources, with what a population can sustain, such as a long duration of highly restrictive measures.

Our modelling suggests that school closure is the least effective single social distancing measure considered, however it is highly disruptive as adults are needed to care for younger children. Its moderate effectiveness arises from our assumption that children still have contact in the wider community when schools are closed. This suggests that combining school closure with even a 30% reduction in community-wide contact will be significantly more effective.

We have evaluated the effectiveness of very robust, combined social distancing interventions, which are very similar to those applied in South Korea (Sophia Rogers, personal communication). Much of the success of the measures used in South Korea is due to the population wishing to work together to minimize infections in others, by complying with the regular guidance given to them by their government *e*.*g*. by daily text messages. It is unclear whether this messaging would be successful in other countries.

Future modelling should evaluate social distancing interventions which might be more applicable in countries such as the UK, other European countries, Australia or the USA. Questions that need answering include which interventions are feasible in a given setting and how effective would these be? Our modelling has assumed that interventions are held until a vaccine or treatment option appears, which may not be feasible, with really strong measures possibly leading to compliance fatigue after being held for long periods of time. To address this, modelling may be used to evaluate repeated cycles of starting and stopping interventions, given that we have demonstrated that starting and stopping robust combined interventions can significantly flatten the epidemic curve.

The COVID-19 transmission characteristics assumed in our model produced an unmitigated infection rate of 66%, which included both symptomatic and asymptomatic cases, and a basic reproduction number of 2.2. Higher or lower R_0_ settings will affect case numbers under all the social distancing interventions considered. In the absence of definitive data on the proportion of infections which are asymptomatic, we assumed a 20% asymptomatic proportion though the actual percentage may be larger. Similarly, if the infectious period is longer than the 3 days assumed, the same caveat applies. However, our sensitivity analyses indicate that changing the underlying model parameters to reflect these modifications does not affect the relative effectiveness of the social distancing measures.

## Data Availability

Not applicable

